# Social dialogue quality and workers’ health as perceived by Belgian trade union representatives during the COVID-19 pandemic

**DOI:** 10.1101/2023.04.10.23288317

**Authors:** Jacques Wels, Natasia Hamarat, Vanessa De Greef

## Abstract

**Background:** Besides major employment disruptions, the COVID-19 pandemic has generated policy responses with specific mechanisms to protect workers’ health. In Belgium, most of these policies were negotiated at national and cross-sectorial level but implemented at company level with company-based collective negotiation playing a key role. This study examines the relationship between trade union representatives’ perception of social dialogue quality and change in workers’ physical and mental health in such a context.

**Methods:** Union representatives were surveyed throughout Belgium between August and December 2021 through an online questionnaire (N=469). We asked about the way they perceived workers’ physical and mental health within their companies and explain variations with the self-perceived change in quality of social dialogue as an exposure. We use a modified Poisson regression for binary outcomes on four stratified models that additively account for no control, company characteristics, pre-pandemic self-reported health and COVID-19-related measures. Weights are generated to ensure sector representativeness.

**Results:** 30.1% of the sample reported a worsening social dialogue quality during the pandemic. Relative Risks (RR) of poor physical and mental health when social dialogue has worsened are respectively 1.49 [95%CI:1.03; 2.15] and 1.38 [95%CI= 1.09;1.74] when controlling for company characteristics and pre-pandemic health. Adding pandemic-related measures reduces the risk of both poor mental [RR=1.25; 95%CI: 0.84; 1.87] and physical health [RR=1.18; 95%CI:0.94;1.49].

**Conclusions:** Although based on self-reported variables, the study shows an association between poor social dialogue quality and poor physical and mental health during the COVID-19 pandemic that must be explored further in post-pandemic context.

## Background

By advocating for workers’ safety, protection of jobs, and fair compensation for those affected by the economic fallout, trade unions have played a crucial role during the COVID-19 pandemic. Yet, the union representatives’ experience of collective negotiation during the pandemic and specifically their role at company level has not been assessed thoroughly. COVID-19 has been seen by many as an occupational disease, the workplace being one of the major sources of infection (Trades Union Congress, 2022). To contain the spread of the virus, state intervention has promoted home working policies like never in the past (Wels et al., 2022) and specific measures have been taken within workplaces where so-called key workers were remaining at work (Watterson, 2020). Similarly, the pandemic has had economic consequences with many countries implementing job retention schemes (i.e., furlough) for part or the totally of their workforce (Wels et al., 2022) and unions playing a role in the implementation and use of these schemes (Müller et al., 2022). In such a context, the capacity for trade unions and employers to carry on with company-level social dialogue has been questioned with some studies showing that the implications of this dialogue have varied depending on pandemic stages (Brandl, 2021) and that unions have, overall, invested in new technologies to pursue collective negotiation (Hunt & Connolly, 2023).

Using a convenience sample collected from union representatives across Belgium, this study aims to better understand how the pandemic might have affected social dialogue quality at company level and how these changes might have translated into different perceptions of workers’ physical and mental health by union representatives. Before expliciting our study’s objectives, we briefly discuss how the relationship between workers’ health and collective negotiation was framed by previous studies. We then describe the specific nature of collective negotiation in Belgium and explain why company-level bargaining is capital to understand the implementation of COVID-19-related policies and their potential effects on workers’ health.

### Collective negotiation and workers’ health

The relationship between employment and population health is well established (Ross & Mirowsky, 1995) but studies on the association between collective negotiation and workers’ health are fewer and mainly focus on three types of approaches.

A first set of studies pays attention to the relationship between union membership and health using cross-sectional, cross-sectorial or macro-level data. For instance, Sochas and Reeves (Sochas & Reeves, 2022) have recently demonstrated using European comparative data that health inequalities are high when unions only represent part of the workforce but low when a high proportion of the workforce is unionized. Similarly, high-country trade union density is associated with lower depressive symptoms among the workforce (Reynolds & Buffel, 2020). The same type of analyses was also made looking at differences across sectors of activity based on union densities (Appleton & Baker, 1985; Taylor, 1987).

A second set of studies has very recently focused on the individual relationship between union membership and health mainly using individual longitudinal data. Results flowing from these studies are quite contradictory showing either a positive (Wels, 2018, 2020) or a negative relationship (Eisenberg-Guyot et al., 2020). A few other studies have focused on the benefits of using a longitudinal approach to assess the association between union membership and wages (Freeman, 1984) or job satisfaction (Bessa et al., 2020) but such a perspective is still rare when looking at health (Wels, 2018, 2020).

Finally, a third set of studies takes a collective approach by focusing on the negotiating process within companies, particularly through health and safety committees. Those committees are set up to negotiate within the workplace working conditions and safety matters and involve trade unions or workers’ representatives. For instance, using Korean cross-sectional data, it was shown that health and safety committees reduce work accidents but seem to be less effective in non-unionized workplaces (Kim & Cho, 2016). By contrast, Bryson has shown for the UK that union representatives within health and safety committees are linked with lower health and safety risks compared with non-unionized workplaces (Bryson, 2016).

The current study takes a collective approach by analysing how union representatives’ perception of social dialogue quality within their company might be associated with how they perceive workers’ health in Belgium during the COVID-19 pandemic. Such a type of study is needed for three reasons. First, using a questionnaire survey, we provide unique quantitative information about the relationship between company level social dialogue quality and workers’ health. No data has ever been collected in Belgium on this matter. Second, we provide evidence of the importance of paying attention to the company-level of negotiation that has been overlooked, which is crucial for an effective policy on wellbeing at work. Thirdly, whilst the relationship between changes in employment settings and workers’ health has been largely documented throughout the pandemic (Adams-Prassl et al., 2020; Hensher, 2020; Topriceanu et al., 2021; Wels et al., 2022), we do not know any quantitative research published on the possible role of collective negotiation in protecting workers during the COVID-19 pandemic and the number of qualitative studies on this matter remains low and mostly theoretical (Franklin, 2021).

### Collective negotiation in Belgium and the role of company-level bargaining

A strong connection exists between worker representation and best practices in health and safety management. The significance of effective worker representation and consultation in improving health and safety outcomes, such as management practices, safety culture, and injury rates was highlighted (Walters, 2010). Key factors for success include a regulatory framework providing rights and facilities for representatives and the means for its enforcement, top management’s commitment to occupational health and safety, and management expertise in hazard and risk assessment and control (Walters et al., 2012; Walters, 2010). Training for representatives and effective communication between worker representatives and their constituents are also vital elements of a successful health and safety committee (Walters & Nichols, 2007). Furthermore, trade unions significantly contribute to the success of worker representation and consultation in health and safety matters (Nichols et al., 2007; Walters, 2010).

In Europe, the EU Framework Directive 89/391 provides that employers shall consult with and permit the participation of workers and/or their representatives in all matters relating to occupational safety and health (OSH).

During the COVID-19 crisis, ILO mentioned that “joint OSH committees have had a critical role to play in responding to OSH concerns [as] they are well positioned to identify situations of potential exposure to COVID-19, assess risks and develop exposure control and mitigation plans” (International Labour Organization, 2022). Legally, joint OSH committees are an important consultative body and several ILO instruments (Convention No. 155 and its Recommendation No. 164) outline the composition and functions of these committees. In Belgium, these OSH Committes - called Committees for Prevention and Protection at Work (CPPT or CPPW – *Comité pour la Prévention et la Protection au Travail* in French, and *Comité voor Preventie en Bescherming op het Werk* in Dutch) - are set up from 50 workers and whose missions are taken over by the trade union delegation or by the workers themselves if the number of 50 is not reached.

Every employer is required to create an internal service for prevention and protection at work, and a worker must serve as a health and safety adviser. If the internal service cannot fulfil all assigned tasks, employers must seek assistance from an external service for prevention and protection at work. In Belgium, an employer without a committee and employing less than twenty workers can himself exercise the function of prevention adviser. The employer, the members of the hierarchy and the occupational health and safety services are involved in the risk assessment and they must consult the Committee’s opinions. On the basis of the risk assessment, the employer shall draw up, in consultation with the members of the line and the occupational health and safety services, an annual and a five-year prevention plan. The Committee must also be consulted on these plans. The employer must regularly evaluate the dynamic risk management system with these actors, which implies in particular taking into account changes in circumstances - like the pandemic - requiring an adaptation of the prevention strategy.

Finally, at intersectoral, sectoral or company level, collective labor agreements may be concluded between one or more representative workers’ organisations and one or more and one or more representative employers’ organisations. During the pandemic, worker health protection was regulated mostly by both intersectoral and sectoral levels to ensure workplace safety and compliance with regulations. For instance, the occupational physicians, who also participate in the CPPT, had their tasks strengthened, as they were mandated to identify high-risk contacts, issue quarantine certificates for high-risk individuals, and to refer workers for testing.

These intentions must be tempered by the use of instruments whose binding force was sometimes not obvious. In that sense, the Higher Council for Prevention and Protection at Work, the *intersectoral* social body in the field of well-being at work, created a generic "guide" to prevent the spread of COVID-19 in the workplace. According to the President of this Council, the guide aimed to address "all companies", including those that had not implemented any measures, often due to a lack of social consultation, and those that had conducted such consultations but implemented inadequate measures (De Greef, 2023). However, the interweaving of the guide with existing standards - which were sometimes more stringent - was not fully thought through and could lead to a less effective system in case of non-compliance.

### COVID-19-related policies and the role of company-level social negotiation

As a result of the many social measures implemented to contain the pandemic many workers have been made redundant (Hensher, 2020), temporary unemployed (Wels et al., 2022) and, for those remaining in employment, home working has gained in importance and is still in place today (Arntz et al., 2020). On the other hand, the workplace has been seen as a major source of COVID-19 exposure with high variations across industries and occupations (Oude Hengel et al., 2022) with particularly high infection rates within the health, education, and public administration sectors in Belgium (Sciensano, 2021).

To support workers and businesses impacted by the pandemic, all applications for temporary unemployment due to force majeure or for economic reasons attributable to the coronavirus, taking effect from 13 March 2020, were automatically being processed under the temporary unemployment scheme due to force majeure. The federal administration adopted a broad interpretation of the notion of force majeure and considered events such as compulsory closure of the shop imposed by the authorities, cancellation of various events (cultural, sports, *etc*.) and absence from work for various reasons to be force majeure (Hachez & De Greef, 2023; Verbruggen, 2020). Starting on the 13^th^ of March 2020, and continuing until the 30^th^ of June 2022, this scheme enabled employers to temporarily halt their workers’ contracts and apply for unemployment benefits on their behalf to prevent job losses and maintain labor market participation. Another example of a job protection measure was the implementation of ‘corona’ paid parental leave from the 1^st^ of May to the 30^th^ of September 2020. After this period had ended, it was replaced with the temporary unemployment scheme, which was available on the 31^st^ of December 2022, for parents with children requiring self-isolation or whose (pre-)school has had to close due to the pandemic.

In 2020, temporary unemployed benefits were granted to almost 1.4 million workers, accounting for over one third of all workers in Belgium (Barrez et al., 2021; Capéau et al., 2022). Alongside the temporary unemployment scheme, the federal government also implemented measures to support self-employed workers who were not eligible for this scheme. For example, a transition allowance (*droit passerelle* / *overbruggingsrecht*) was extended several times - by relaxing the conditions - for the self-employed workers who had to stop their activity due to the pandemic (Detienne, 2022). In June 2020, it was decided that self-employed persons who are obliged to stop or reduce their activities until at least the beginning of May could continue to benefit from this allowance if after full resumption of their activities provided that they could demonstrate that the activity had experienced a decrease of at least 10% in turnover or orders compared to a defined period (Detienne, 2022). They could also apply for exemption or partial remission of the payment of social security contributions. In addition to these measures, various compensation mechanisms for businesses and self-employed individuals who had to stop their activities due to the lockdown measures were implemented at regional level (Capéau et al., 2022).

At the onset of the pandemic, Belgium experienced a sudden increase in the number of people doing home working, much like many other countries. From April 3^rd^ to May 3^rd^, 2020, teleworking at home becomes mandatory in so-called *non-essential sectors*. This was later highly recommended until the 2^nd^ of November 2020, when it became mandatory for the second time, unless this is impossible due to the nature of the function, the continuity of the management of the company, its activities or its services. This obligation was lifted on the 27^th^ of June 2021, although home working remained strongly recommended until the 31^st^ of August 2021. Starting from the 29^th^ of October 2021, teleworking at home was again strongly recommended, and it became mandatory for the third time from the 20^th^ of November 2021, except for one day a week from the 26^th^ of December 2021 to the 18^th^ of February 2022 (ensuring that a maximum of 20% of those for whom telework at home is mandatory are present at the workplace at the same time). From that point on, home working remains recommended until March 7, 2022.

To address these changes, the Belgian federal government implemented a few measures to support telework at home and ensure the health and safety of workers. One of these was the reimbursement of expenses incurred due to telework at home, including the purchase of office equipment, and regular expenses such as heating and electricity. For example, the employer could intervene in a lump sum by paying a monthly allowance of up to 129.48 euros per month to workers who work from home. The allowance then increased to a maximum of 144.31 euros per month for the months of April to September 2021, rising to 145.81 euros from the 1^st^ of December 2022. This allowance was considered not as taxable remuneration and was not subject to social security contributions. Each employer was invited to define their conditions during the pandemic, and the social partners within the National Labour Council concluded a collective labour agreement on the 26^th^ of January 2021, establishing obligations and recommendations for employers who had not yet established agreements on home working. In addition, the Belgian federal government introduced measures to support workers who were unable to work from home, such as those in essential sectors^1^. Federal and regional levels introduced various financial support for employers to help cover the costs of additional protective measures, such as personal protective equipment, but also incentives (e.g. incentives for nurses and assistant nurses).

All these measures reveal the policies put in place but do not allow us to understand the negotiation at company level. The outcome of social dialogue at company level is unclear, not only because it naturally varies by company, but also because the agreements that result from this negotiation are inaccessible for research. For this reason, we established a questionnaire to understand what companies had concretely established to maintain social dialogue, while wanting to know more about the health of workers in these companies. Specifically, the aim of this study is to measure the impact of social dialogue on workers’ health at the company level and to contribute to the literature that attempts to measure the effects of social concertation.

### Study’s objectives

The primary objective of this study is to assess how company-level social dialogue quality has changed after the start of the COVID-19 pandemic and whether this change translates into a change in perception of workers’ physical and mental health by trade union representatives. Two sub-research questions are investigated. First, we assess whether such a relationship is still effective after successively controlling for company characteristics, self-reported pre-pandemic health outcomes and policy and company measures implemented during the pandemic. Second, we examine whether pandemic-related policy responses such as furlough, home working or specific workers’ health protection measures implemented at state level are associated with a change in social dialogue quality.

## Data and methods

### Data collection

Due to lack data on both union representative’s perception of workers’ health and social dialogue, data were collected over a five-month span between the 12^th^ of August 2021 and the 20^th^ of December 2021 through an online questionnaire (in French and Dutch). Contacts have been made with the intersectoral negotiating bodies (National Labor Council and the Superior Council for Prevention and Protection at Work). Unfortunately, very few employers’ representatives responded to the questionnaire but it was shared widely through the main Belgian trade unions (N=469).

### Outcome variable: perceived workers’ health

It was asked to respondents how they would qualify the physical and mental health (as two separated questions) of their company’s workers since the start of the COVID-19 crisis (March 2020) with five possible answers: (1) very good health, (2) good health, (3) average health, (4) poor health and (5) very poor health. The same question was replicated later about pre-pandemic physical and mental health. To facilitate the interpretation of the variable, we have generated a new dummy variable that combines ‘average’, ‘good’ and ‘very good’ into one reference modality (0) and ‘poor’ and ‘very poor’ as another modality (1). Among the full set of respondents (N=469), 103 declared a poor to very poor physical health (against 366 who declared an average to very good physical health) and 206 declared a poor to very poor mental health (against 263 who declared an average to very good physical health).

### Exposure: Quality of Social dialogue

The quality of social dialogue is measured as the response to one single item asking respondents how they would qualify the quality of social dialogue with three possible modalities: (1) better than before the COVID-19 crisis, (2) same as before the COVID-19 crisis, and (3) worse than before the COVID-19 crisis. Non-weighted descriptive data show that 257 respondents declared that the quality of social dialogue was the same, 33 declared that it was better, 141 declared that it was worst and 38 did not know.

### Covariates

The study controls for a list of covariates including:

– The *share of men and women* working within the company (half women, mainly women, mainly men (reference));
– the *average age* of workers (mainly higher than 40, about 40, mainly lower than 40 (reference));
– the *size of the company* (>1,200; 600-1200; 300-599; 100-299; 50-99; <49 (reference));
– the *self-qualified type of work* within the company (intellectual, manual, both (reference));
– the *questionnaire’s language* (Dutch, French (reference)); the *Joint Committee* (200-299, 300-399, None, Public sector, 100-199 (reference));
– *pre-pandemic mental health,* whether a *new collective agreement* was concluded during the COVID-19 pandemic (no, do not know, yes (reference));
– the *role of the health and safety adviser* during the COVID-19 pandemic (Good, poor, average (reference));
– whether a *risk assessment* was made during the COVID-19 pandemic (No, yes (reference));
– how many employees were *working from home* during the first and second lockdowns (two separated variables) (50 percent or lower; 75 to 100 percent; between 50 and 75 percent (reference));
– whether *a redundancy plan* was implemented during the COVID-19 pandemic (yes, no (reference));
– whether part of the workforce was made *furlough* during the first and second lockdowns (two separated variables) (Yes, no (reference));
– whether the sector is self-perceived as critical (*key worker*) (yes, no (reference)).

### Adjustment levels

We use four additive levels of adjustments. First, we look at the association between change in social dialogue and self-reported workers’ physical/mental health without control variables (*No adjustment*). Second, we include company’s characteristics such as gender ratio, age ratio, company size, language, type of work and joint committee (*company adjustment*). Third, we include pre-pandemic self-reported health information (*Pre-pandemic health adjustment*). Finally, we control for the full set of covariates including whether a new agreement was made, whether a risk assessment took place, the share of employees working from home and furloughed, whether a redundancy plan was started and whether workers categorised themselves as key workers (*Full adjustment*).

### Modified Poisson regression

We use Modified Poisson Regression for binary outcomes with robust standard errors (sandwich estimator) (Zou, 2004; Zou & Donner, 2013) that allows calculating the Relative Risks instead of the Odds Ratios (Ranganathan et al., 2015).

### Normalised sector Weight

Data were weighted using a normalized sector weight aiming to adjust Joint Committee representativeness using full population data from the Datawarehouse Marché du Travail et Protection Sociale (**table 1**).

**Table 1.** Sample distribution by joint commission and actual distribution.

|  | **Survey** |  | **DWMTPS 2020/4\*** |  |
| --- | --- | --- | --- | --- |
|  | *N* | *Per cent* | *N* | *Per cent* |
| CP 100-199 | 78 | 16.7 | 671,371 | 18.7 |
| CP 200-299 | 150 | 31.9 | 832,473 | 23.2 |
| CP 300-399 | 171 | 36.4 | 1,144,461 | 31.9 |
| Public sector<br>and no CP | 70 | 14.9 | 942,155 | 26.2 |
| Total | 494 | 100 | 3,590,460 | 100 |

Source: \* Datawarehouse Marché du Travail et Protection Sociale, 4<sup>th</sup> semester of 2020

## Results

### Descriptive statistics

Table 1 exhibits the distribution of the sample across 4 levels of Joint Committees (CPs) and compares this distribution with the actual distribution within the full employed workforce in 2020. It can be observed that the distribution within the survey is not fully different with the distribution of the employed workforce within the whole population with 16.7 percent of the sample in the CP 100 to 199 in the survey against 18.7 in the population. CP 200 to 299 and 300 to 399 are slightly overrepresented with, respectively, 31.9 and 36.4 percent of the sample against 23.2 and 31.9 within the population. This is explained by a lower presence of public sector respondents or representatives with no Joint Commission: 14.9 in our sample against 26.2 in the population. To tackle this representativeness bias, we use a normalised population weight.

Further descriptive statistics are provided in <u>supplementary file 1</u>. Looking at weighted percentages, it can be observed that poor self-perceived workers’ physical health was 3.4 percent prior the start of the pandemic and 21.1 percent during the pandemic, an increase of 17.7 percentage points. Similarly, poor self-perceived workers’ mental health 8.8 percent against 42.8 percent during the pandemic, an increase of 34 percentage points. Not surprisingly, workers’ mental health is perceived to have deteriorated during the pandemic. The quality of social dialogue has also changed. 53.1 percent of the sample reports a social dialogue quality that remained the same since the pandemic’s outbreak. 7.8 percent of the weighted sample reports an improved social quality dialogue against 30.1 percent who reported a deteriorated social dialogue within their company. 9 percent of the sample do not know whether social dialogue has changed.

Other variables on company characteristics show the diversity of the companies represented within the sample. 44 per cent of the union representatives qualified their company as doing mainly an intellectual work against 14.6 where work was qualified as manual. 41.5 percent of the respondents described it as both manual and intellectual. Looking at sex, respectively 19.3 and 25.1 percent of the sample reported a workforce that was mainly composed of male or female. 55.1% reported a mixed sex composition. The same kind of distribution is found when looking at the estimated mean age of the company’s workers with 21 percent reporting a mean age below 40, 34.5 percent reporting a mean age above 40 and 44.5 percent reporting a mean age of 40. Most companies included in the study have a size that is over 50 as only 9.8 percent of the sample reported working in a company that employs less than 50 workers.

Finally, looking at the measures implemented during the pandemic, we observe that only 37.6 percent the workforce’s representatives declared that a risk evaluation was made against 35.6 percent who declared that it was not made. Interestingly, 26.8 per cent of the sample do not know whether a risk evaluation was made. About half (45 per cent) of the representatives mentioned that furlough was implemented within their company after the first lockdown and 35.4 percent after the second lockdown. Home working^2^ was also represented with different schemes throughout the pandemic. 64.9 percent of the sample reported that they were working in a key sector of activity. This is not surprising as it is known that self-reported key sector is likely to be higher than a key sector definition that would be based on sector of activity (Wielgoszewska et al., 2022).

### Estimates

Estimates for physical health as an outcome are shown in **table 2**.

**Table 2.**
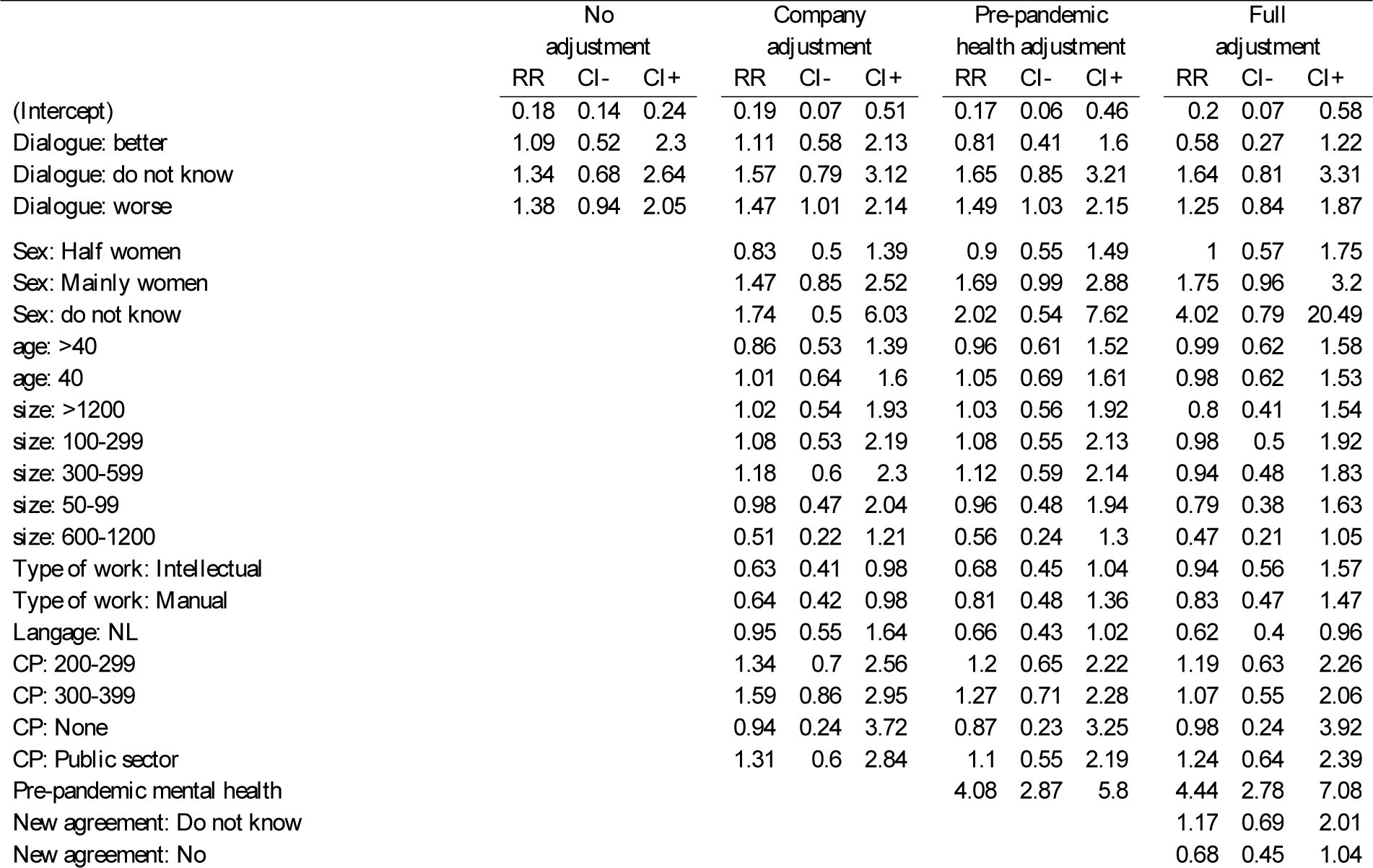
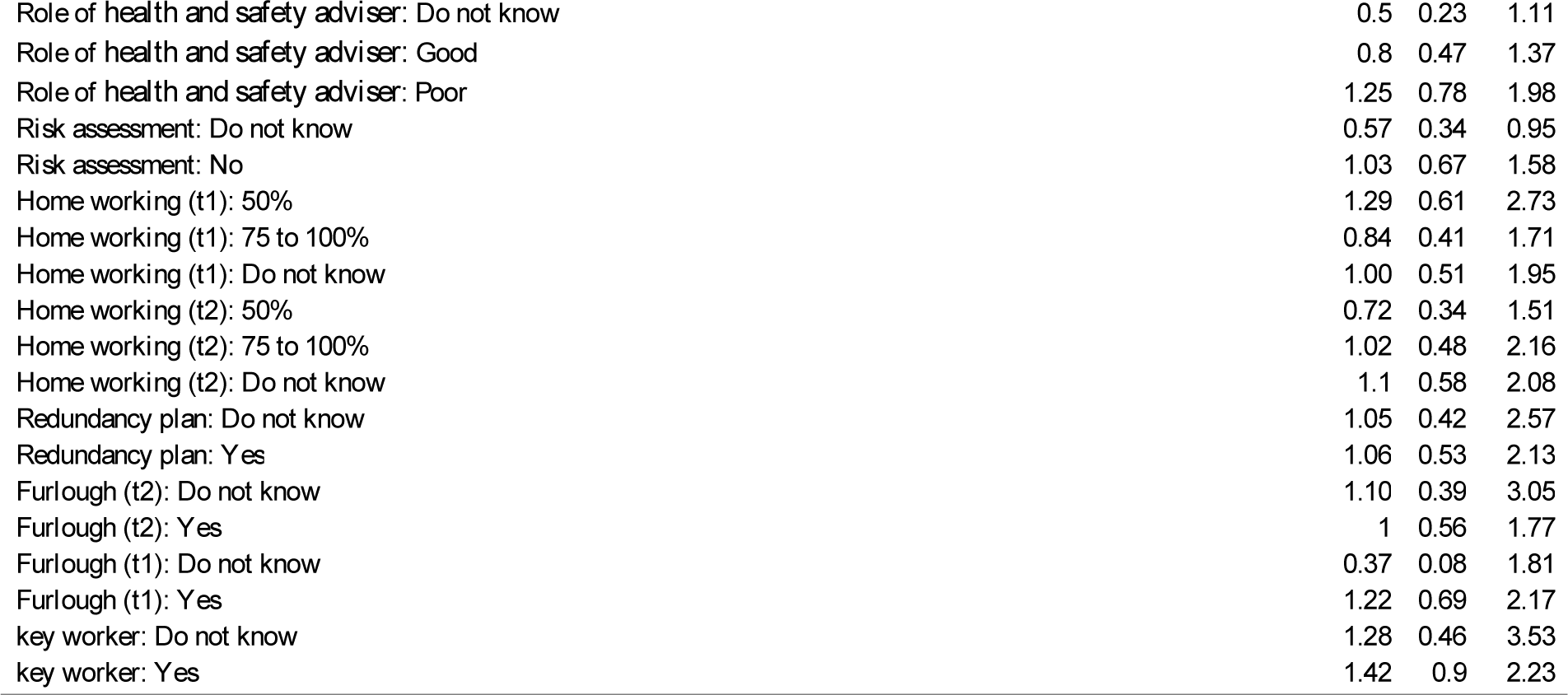
Association between self-reported workers’ physical health and social dialogue. Modified Poisson Regression with Relative Risks and 95% confidence interval.

|  | No adjustment |  |  | Company adjustment |  |  | Pre-pandemic health adjustment |  |  | Full adjustment |  |  |
| --- | --- | --- | --- | --- | --- | --- | --- | --- | --- | --- | --- | --- |
|  | RR | CI- | CI+ | RR | CI- | CI+ | RR | CI- | CI+ | RR | CI- | CI+ |
| (Intercept) | 0.18 | 0.14 | 0.24 | 0.19 | 0.07 | 0.51 | 0.17 | 0.06 | 0.46 | 0.2 | 0.07 | 0.58 |
| Dialogue: better | 1.09 | 0.52 | 2.3 | 1.11 | 0.58 | 2.13 | 0.81 | 0.41 | 1.6 | 0.58 | 0.27 | 1.22 |
| Dialogue: do not know | 1.34 | 0.68 | 2.64 | 1.57 | 0.79 | 3.12 | 1.65 | 0.85 | 3.21 | 1.64 | 0.81 | 3.31 |
| Dialogue: worse | 1.38 | 0.94 | 2.05 | 1.47 | 1.01 | 2.14 | 1.49 | 1.03 | 2.15 | 1.25 | 0.84 | 1.87 |
| Sex: Half women |  |  |  | 0.83 | 0.5 | 1.39 | 0.9 | 0.55 | 1.49 | 1 | 0.57 | 1.75 |
| Sex: Mainly women |  |  |  | 1.47 | 0.85 | 2.52 | 1.69 | 0.99 | 2.88 | 1.75 | 0.96 | 3.2 |
| Sex: do not know |  |  |  | 1.74 | 0.5 | 6.03 | 2.02 | 0.54 | 7.62 | 4.02 | 0.79 | 20.49 |
| age: >40 |  |  |  | 0.86 | 0.53 | 1.39 | 0.96 | 0.61 | 1.52 | 0.99 | 0.62 | 1.58 |
| age: 40 |  |  |  | 1.01 | 0.64 | 1.6 | 1.05 | 0.69 | 1.61 | 0.98 | 0.62 | 1.53 |
| size: >1200 |  |  |  | 1.02 | 0.54 | 1.93 | 1.03 | 0.56 | 1.92 | 0.8 | 0.41 | 1.54 |
| size: 100-299 |  |  |  | 1.08 | 0.53 | 2.19 | 1.08 | 0.55 | 2.13 | 0.98 | 0.5 | 1.92 |
| size: 300-599 |  |  |  | 1.18 | 0.6 | 2.3 | 1.12 | 0.59 | 2.14 | 0.94 | 0.48 | 1.83 |
| size: 50-99 |  |  |  | 0.98 | 0.47 | 2.04 | 0.96 | 0.48 | 1.94 | 0.79 | 0.38 | 1.63 |
| size: 600-1200 |  |  |  | 0.51 | 0.22 | 1.21 | 0.56 | 0.24 | 1.3 | 0.47 | 0.21 | 1.05 |
| Type of work: Intellectual |  |  |  | 0.63 | 0.41 | 0.98 | 0.68 | 0.45 | 1.04 | 0.94 | 0.56 | 1.57 |
| Type of work: Manual |  |  |  | 0.64 | 0.42 | 0.98 | 0.81 | 0.48 | 1.36 | 0.83 | 0.47 | 1.47 |
| Langage: NL |  |  |  | 0.95 | 0.55 | 1.64 | 0.66 | 0.43 | 1.02 | 0.62 | 0.4 | 0.96 |
| CP: 200-299 |  |  |  | 1.34 | 0.7 | 2.56 | 1.2 | 0.65 | 2.22 | 1.19 | 0.63 | 2.26 |
| CP: 300-399 |  |  |  | 1.59 | 0.86 | 2.95 | 1.27 | 0.71 | 2.28 | 1.07 | 0.55 | 2.06 |
| CP: None |  |  |  | 0.94 | 0.24 | 3.72 | 0.87 | 0.23 | 3.25 | 0.98 | 0.24 | 3.92 |
| CP: Public sector |  |  |  | 1.31 | 0.6 | 2.84 | 1.1 | 0.55 | 2.19 | 1.24 | 0.64 | 2.39 |
| Pre-pandemic mental health |  |  |  |  |  |  | 4.08 | 2.87 | 5.8 | 4.44 | 2.78 | 7.08 |
| New agreement: Do not know |  |  |  |  |  |  |  |  |  | 1.17 | 0.69 | 2.01 |
| New agreement: No |  |  |  |  |  |  |  |  |  | 0.68 | 0.45 | 1.04 |

|  |  |  |  |
| --- | --- | --- | --- |
| Role of health and safety adviser: Do not know | 0.5 | 0.23 | 1.11 |
| Role of health and safety adviser: Good | 0.8 | 0.47 | 1.37 |
| Role of health and safety adviser: Poor | 1.25 | 0.78 | 1.98 |
| Risk assessment: Do not know | 0.57 | 0.34 | 0.95 |
| Risk assessment: No | 1.03 | 0.67 | 1.58 |
| Home working (t1): 50% | 1.29 | 0.61 | 2.73 |
| Home working (t1): 75 to 100% | 0.84 | 0.41 | 1.71 |
| Home working (t1): Do not know | 1.00 | 0.51 | 1.95 |
| Home working (t2): 50% | 0.72 | 0.34 | 1.51 |
| Home working (t2): 75 to 100% | 1.02 | 0.48 | 2.16 |
| Home working (t2): Do not know | 1.1 | 0.58 | 2.08 |
| Redundancy plan: Do not know | 1.05 | 0.42 | 2.57 |
| Redundancy plan: Yes | 1.06 | 0.53 | 2.13 |
| Furlough (t2): Do not know | 1.10 | 0.39 | 3.05 |
| Furlough (t2): Yes | 1 | 0.56 | 1.77 |
| Furlough (t1): Do not know | 0.37 | 0.08 | 1.81 |
| Furlough (t1): Yes | 1.22 | 0.69 | 2.17 |
| key worker: Do not know | 1.28 | 0.46 | 3.53 |
| key worker: Yes | 1.42 | 0.9 | 2.23 |

Looking at the quality of social dialogue in the non-adjusted model, we observe that neither perceiving a worse dialogue nor a better dialogue compared to pre-pandemic time is significantly associated with poorer self-reported health outcomes. In other words, the relative risk of poor self-reported health is 1.38 [95%CI= 0.94; 2.05] when the change in social dialogue is perceived negatively and 1.09 [95%CI= 0.52; 2.30] when it is seen positively.

Estimates are quite different when adjusting for company’s characteristics (company adjustment) and pre-pandemic health (pre-pandemic health adjustment). It can be observed that the relationship between reporting a worsening social dialogue and the relative risk of perceived a poor self-reported workers’ health is positive and significant: 1.47 (95%CI= 1.01; 2.14) after adjusting for company’s characteristics and 1.49 [95%CI= 1.03; 2.15] after adjusting for pre-pandemic physical health. In other words, these two levels of adjustment show a positive and significant association between reporting a worse social dialogue and reporting a poor perceived workers’ health. No significant relationship is observed for the covariates except for the type of work and pre-pandemic reported health. Describing work as intellectual or manual (versus perceiving the work as mixed) is significantly associated with lower risk of poor physical health in the model adjusted for company characteristics with relative risks of respectively 0.63 [95%CI= 0.41; 0.98] and 0.64 [95%CI= 0.42; 0.98] but this significant relationship fades away after controlling for pre-pandemic physical health which translates a difference that existed prior the start of the pandemic.

Controlling for COVID-19-related measures drastically reduces the significance of such a relationship. The fourth adjustment mode shows a relative risk for those reporting a worse social dialogue of 1.25 [95%CI= 0.85; 1.87], which overlaps the null hypothesis. However, no significant coefficient is observed for the COVID-19-related covariates.

Analyses were replicated using self-reported workers’ mental health as an outcome. Estimates are of the same nature but with both higher intensities and smaller confidence intervals, indicating a stronger perceived change in mental health during the pandemic, as documented by previous studies for the global population (Patel et al., 2022) as well as for the workforce (Wels & Hamarat, 2021). Results in **table 3** show a positive and statistically significant association between reporting a worse social dialogue and a poorer self-reported perceived physical health of the workers (RR:1.40, 95%CI= 1.11; 1.78) in the non-adjusted model. This relationship stands when company-related variables and pre-pandemic mental health are introduced in the model with relative risks of respectively 1.41 (95%CI= 1.12; 1.79) and 1.38 (95%CI= 1.09; 1.74). As for the physical health model, controlling for COVID-19-related measures reduces the intensity of the relationship (RR=1.18) and broadens the confidence interval (95%CI= 0.94, 1.49). Three covariates are associated with a poorer self-reported workers’ health. The size of the company, and particularly working in a company of more than 1,200 workers, is associated with a poorer self-reported health (RR:1.77, 95%CI= 1.13, 2.78 in the fully adjusted model). Pre-pandemic mental health is strongly associated with pandemic variable with a risk of 2.49 (95%CI= 2.03, 3.05) in the fully adjusted model. Finally, a poor role of the health and safety adviser is associated with poor mental health outcomes for the workforce (RR:1.67, 95%CI= 1.27, 2.20), which was not observed when looking at physical health as an outcome.

**Table 3.**
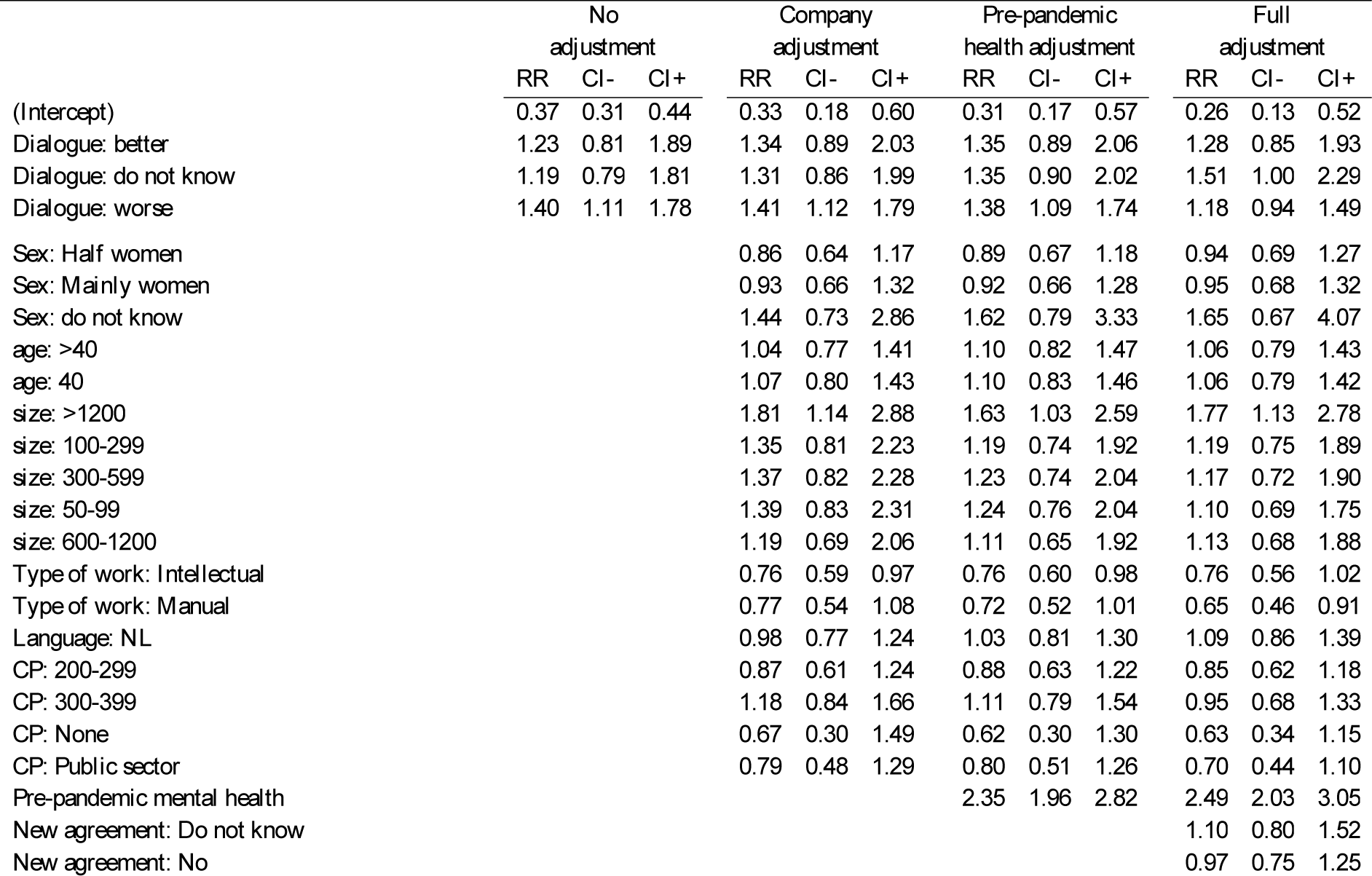
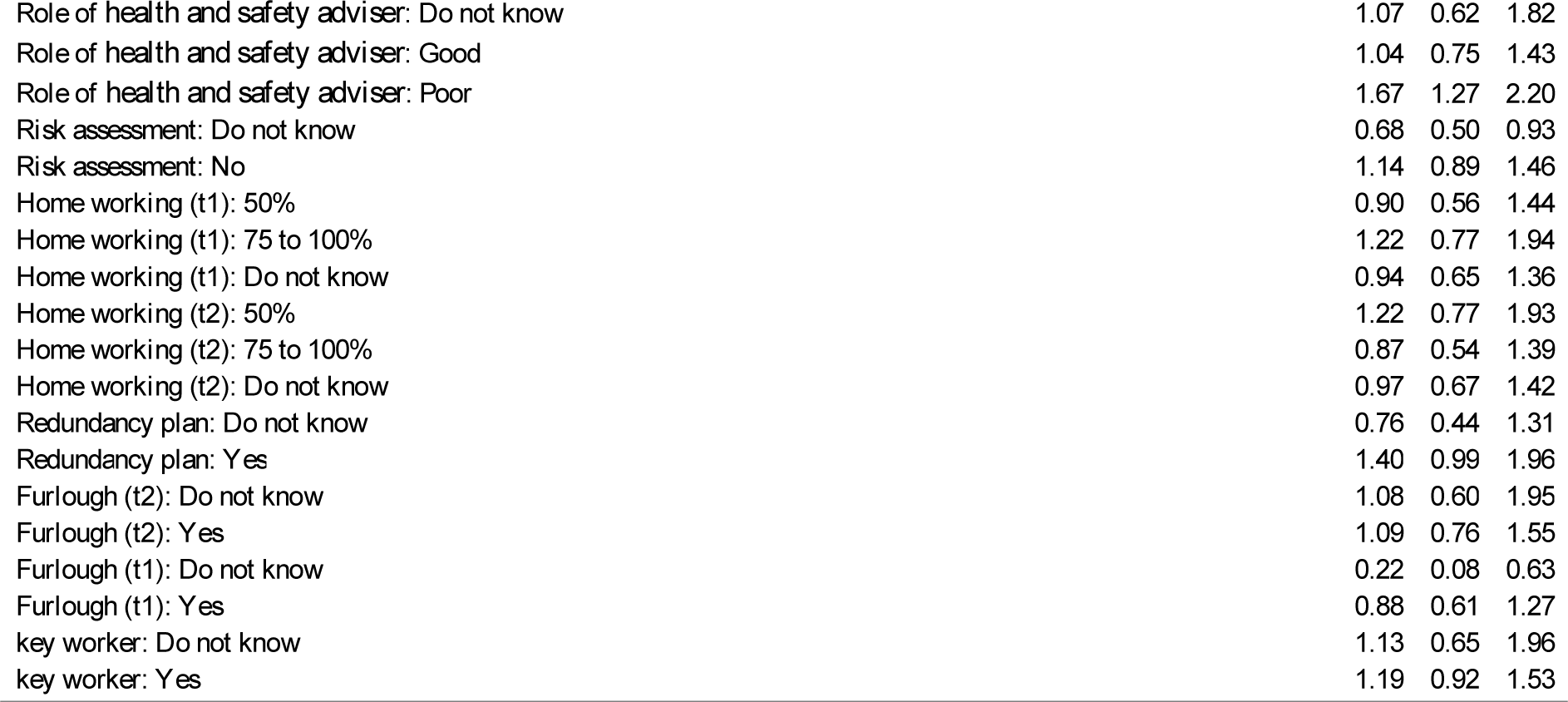
Association between self-reported workers’ mental health and social dialogue. Modified Poisson Regression with Relative Risks and 95% confidence interval

|  | No adjustment |  |  | Company adjustment |  |  | Pre-pandemic health adjustment |  |  | Full adjustment |  |  |
| --- | --- | --- | --- | --- | --- | --- | --- | --- | --- | --- | --- | --- |
|  | RR | CI- | CI+ | RR | CI- | CI+ | RR | CI- | CI+ | RR | CI- | CI+ |
| (Intercept) | 0.37 | 0.31 | 0.44 | 0.33 | 0.18 | 0.60 | 0.31 | 0.17 | 0.57 | 0.26 | 0.13 | 0.52 |
| Dialogue: better | 1.23 | 0.81 | 1.89 | 1.34 | 0.89 | 2.03 | 1.35 | 0.89 | 2.06 | 1.28 | 0.85 | 1.93 |
| Dialogue: do not know | 1.19 | 0.79 | 1.81 | 1.31 | 0.86 | 1.99 | 1.35 | 0.90 | 2.02 | 1.51 | 1.00 | 2.29 |
| Dialogue: worse | 1.40 | 1.11 | 1.78 | 1.41 | 1.12 | 1.79 | 1.38 | 1.09 | 1.74 | 1.18 | 0.94 | 1.49 |
| Sex: Half women |  |  |  | 0.86 | 0.64 | 1.17 | 0.89 | 0.67 | 1.18 | 0.94 | 0.69 | 1.27 |
| Sex: Mainly women |  |  |  | 0.93 | 0.66 | 1.32 | 0.92 | 0.66 | 1.28 | 0.95 | 0.68 | 1.32 |
| Sex: do not know |  |  |  | 1.44 | 0.73 | 2.86 | 1.62 | 0.79 | 3.33 | 1.65 | 0.67 | 4.07 |
| age: >40 |  |  |  | 1.04 | 0.77 | 1.41 | 1.10 | 0.82 | 1.47 | 1.06 | 0.79 | 1.43 |
| age: 40 |  |  |  | 1.07 | 0.80 | 1.43 | 1.10 | 0.83 | 1.46 | 1.06 | 0.79 | 1.42 |
| size: >1200 |  |  |  | 1.81 | 1.14 | 2.88 | 1.63 | 1.03 | 2.59 | 1.77 | 1.13 | 2.78 |
| size: 100-299 |  |  |  | 1.35 | 0.81 | 2.23 | 1.19 | 0.74 | 1.92 | 1.19 | 0.75 | 1.89 |
| size: 300-599 |  |  |  | 1.37 | 0.82 | 2.28 | 1.23 | 0.74 | 2.04 | 1.17 | 0.72 | 1.90 |
| size: 50-99 |  |  |  | 1.39 | 0.83 | 2.31 | 1.24 | 0.76 | 2.04 | 1.10 | 0.69 | 1.75 |
| size: 600-1200 |  |  |  | 1.19 | 0.69 | 2.06 | 1.11 | 0.65 | 1.92 | 1.13 | 0.68 | 1.88 |
| Type of work: Intellectual |  |  |  | 0.76 | 0.59 | 0.97 | 0.76 | 0.60 | 0.98 | 0.76 | 0.56 | 1.02 |
| Type of work: Manual |  |  |  | 0.77 | 0.54 | 1.08 | 0.72 | 0.52 | 1.01 | 0.65 | 0.46 | 0.91 |
| Language: NL |  |  |  | 0.98 | 0.77 | 1.24 | 1.03 | 0.81 | 1.30 | 1.09 | 0.86 | 1.39 |
| CP: 200-299 |  |  |  | 0.87 | 0.61 | 1.24 | 0.88 | 0.63 | 1.22 | 0.85 | 0.62 | 1.18 |
| CP: 300-399 |  |  |  | 1.18 | 0.84 | 1.66 | 1.11 | 0.79 | 1.54 | 0.95 | 0.68 | 1.33 |
| CP: None |  |  |  | 0.67 | 0.30 | 1.49 | 0.62 | 0.30 | 1.30 | 0.63 | 0.34 | 1.15 |
| CP: Public sector |  |  |  | 0.79 | 0.48 | 1.29 | 0.80 | 0.51 | 1.26 | 0.70 | 0.44 | 1.10 |
| Pre-pandemic mental health |  |  |  |  |  |  | 2.35 | 1.96 | 2.82 | 2.49 | 2.03 | 3.05 |
| New agreement: Do not know |  |  |  |  |  |  |  |  |  | 1.10 | 0.80 | 1.52 |
| New agreement: No |  |  |  |  |  |  |  |  |  | 0.97 | 0.75 | 1.25 |

|  |  |  |  |
| --- | --- | --- | --- |
| Role of health and safety adviser: Do not know | 1.07 | 0.62 | 1.82 |
| Role of health and safety adviser: Good | 1.04 | 0.75 | 1.43 |
| Role of health and safety adviser: Poor | 1.67 | 1.27 | 2.20 |
| Risk assessment: Do not know | 0.68 | 0.50 | 0.93 |
| Risk assessment: No | 1.14 | 0.89 | 1.46 |
| Home working (t1): 50% | 0.90 | 0.56 | 1.44 |
| Home working (t1): 75 to 100% | 1.22 | 0.77 | 1.94 |
| Home working (t1): Do not know | 0.94 | 0.65 | 1.36 |
| Home working (t2): 50% | 1.22 | 0.77 | 1.93 |
| Home working (t2): 75 to 100% | 0.87 | 0.54 | 1.39 |
| Home working (t2): Do not know | 0.97 | 0.67 | 1.42 |
| Redundancy plan: Do not know | 0.76 | 0.44 | 1.31 |
| Redundancy plan: Yes | 1.40 | 0.99 | 1.96 |
| Furlough (t2): Do not know | 1.08 | 0.60 | 1.95 |
| Furlough (t2): Yes | 1.09 | 0.76 | 1.55 |
| Furlough (t1): Do not know | 0.22 | 0.08 | 0.63 |
| Furlough (t1): Yes | 0.88 | 0.61 | 1.27 |
| key worker: Do not know | 1.13 | 0.65 | 1.96 |
| key worker: Yes | 1.19 | 0.92 | 1.53 |

Analyses were replicated using a binary logit regression (providing odds ratios – OR) instead of a modified Poisson (providing relative risks – RR). Estimates for the variable of interest for each level of adjustment are shown in **figure 1** with no difference in the direction of the estimates but stronger effects for the binary logit model showing that odds ratios are exaggerated when the outcome is common, giving the impression of a large effect when the outcome is common.

**Figure 1.**
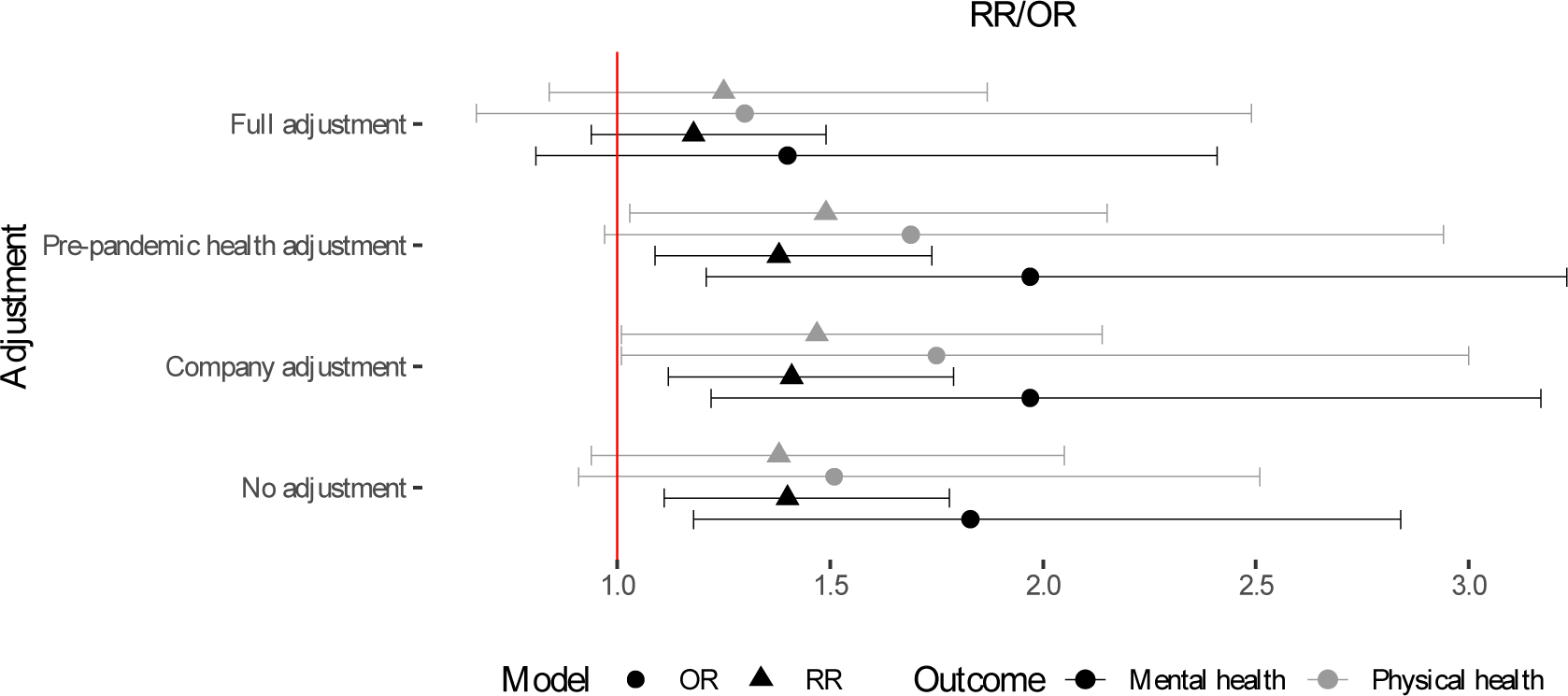
Comparison between Relative Risks and Odds Ratios.

Additional non-weighted models are replicated in supplementary file 2 with no major differences across estimates.

Previous results show that the relationship between the poor quality of social dialogue and poor physical and mental health is strong and significant when controlling for companies’ characteristics and pre-pandemic mental and physical health. Although still positive, such an association is not significant after introducing COVID-19-related measures such as home working, furlough or whether a risk assessment was made. One issue is that those changes might be someway related to the quality of the social dialogue. To assess these associations, we performed a Chi-square test looking at the relationship between the change in social dialogue and whether a new collective agreement was concluded during the COVID-19 pandemic, whether furlough was used within the company, the share of the workforce that was working remotely, whether the company has implemented a redundancy plan, whether a risk evaluation was made, whether the respondent reported working in a key sector of activity and the self-evaluated role of the health and safety adviser during the pandemic. Results of the Chi-square test are shown in **figure 2**.

**Figure 2.**
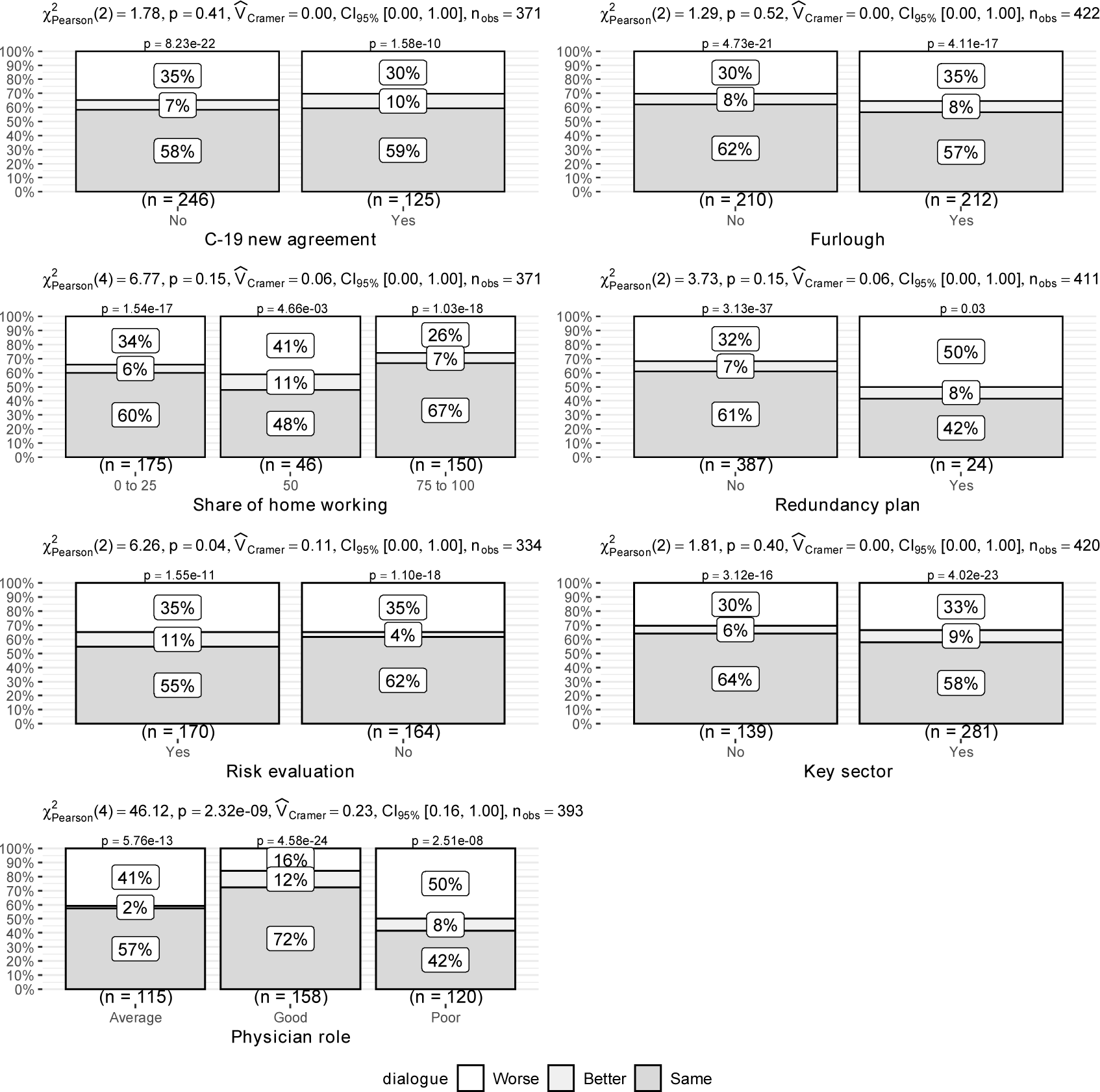
Chi-square tests.

Neither a new collective agreement, the use of furlough, the share of workers working remotely nor whether workers were part of a key sector of activity are associated with a change in social dialogue quality. Plots do not show stronger discrepancies and results of the Chi-square test are not significant. The implementation of a redundancy plan by the employer only concerns a minority of respondents (N=24) and is associated with poorer social dialogue for 50% of them. However, this association should be taken with caution because redundancy plans in Belgium are implemented at employers’ discretion, on the one hand, and because the association only applies to a very limited sample, providing an association that is not statistically significant, on the other hand. Only the presence of a risk assessment and the role of the health and safety adviser is significantly associated with a change in social dialogue quality. In companies where a risk assessment was made, 11 per cent of the representatives reported a better workers’ health against 4 per cent in companies where no risk assessment was made. Results indicate that a risk assessment is associated with an increase in the percentage of respondents reporting a better social dialogue but without reducing the percentage of those reporting a poorer social dialogue. By contrast, the role of the health and safety adviser is clearly and significantly associated with the quality of social dialogue. In companies where the role of the health and safety adviser is assessed as poor, 50 per cent of the respondents reported a poorer workforce health against 41 per cent when their role is perceived as average and 16 per cent when it is perceived as good.

## Discussion

The COVID-19 pandemic has generated many employment disruptions and has challenged our occupational health prevention systems. Several studies have pointed out the deep connection between changes in work and employment settings throughout the pandemic and workers physical and mental health. Attention has been given to the increased number of workers who have been working from home, on temporary unemployment schemes (furlough) or have lost their job during the pandemic but studies looking at the way collective bargaining and social dialogue could have contributed to improving workers’ mental and physical health are nearly inexistent.

Our study brings a unique overview on such a relationship. Four key findings could be pointed out. First, self-reported workers’ health as perceived by union representatives is seen to have declined during the pandemic. This is particularly true for mental health but also for physical health and echoes what has been found using panel data in other countries (Di Gessa et al., 2021; Patel et al., 2022). Second, the pandemic is perceived to have slightly affected the quality of social dialogue within Belgian companies. Whilst more than half of our sample (54.8 percent) reports that social dialogue quality has not changed, 30.1% of the respondents has reported that social dialogue quality has worsened during the pandemic. Third, the study shows an association between social dialogue quality and perceived workers’ mental and physical health that is independent of company characteristics and pre-pandemic health. Relative risks are higher when looking at mental health but a relationship with physical health is also observed. Finally, we observe that the associations remain positive when controlling for some pandemic-related measures (i.e., home working, furlough, redundancy plan, key sector, risk assessment, new collective agreement, role of the health safety adviser) but relative risks are lower and not significant at 95 percent compared with previous levels of adjustment. There is an association between social dialogue quality and the role of the physician or health and safety adviser and whether a risk assessment was made, which may indicate that these two variables are somehow connected with social dialogue quality.

Whilst this study brings unique insights on the relationship between social dialogue and occupational health at company level in Belgium where no systematic data collection takes place, this study is not without limitations. The major pitfall is that it mainly relies on union representatives’ point of view on both change in social dialogue quality and the physical and mental health of the workforce they represent. Subjective health for oneself is an accurate indicator of general health and an accurate predicator of both mental and physical health (Ploubidis & Grundy, 2011; Ploubidis & Pongiglione, 2019) but the health of a group of people as reported by only one individual might be affected by both individual and exogenous factors. Similarly, the study is not longitudinal and calculates measures of the change in health and social dialogue that are based on retrospective information and not on pre-pandemic collected data (although the timing of collection of pre-pandemic variables might also be an issue (Wels, et al., 2022)) and could not be linked with other administrative data or surveys. As no other Belgian study specifically focuses on both health and social dialogue and no question is specifically asked to union representatives, the only possible way to answer our research questions was to collect our own data. Focusing on another country – where the collective negotiation process as well as the measures implemented during the COVID-19 pandemic might be quite distinct – would not have possibly allowed to answer those research questions. Whilst the use of an online questionnaire should be avoided to provide adequate responses to these research questions, particularly due to sampling problems and low response rates, we have attempted to maximise response rates and have used sector-based weights to correct the sample in the most efficient way. Finally, it has to be mentioned that despite our attempts to collect information about the way employers and employers’ representatives have perceived social dialogue and workers’ health during the pandemic, we obtained very low response rates on the employers’ side (<5) and had to focus on the workers’ representatives’ side only for the quantitative part of our research.

From a more theoretical perspective, our research also discusses three aspects. First, we demonstrate that company-level negotiation plays a pivotal role in implementing sectorial, cross-sectorial and national policies. Unlike countries such as the United Kingdom where companies became the central level of collective negotiation or Japan where sectorial negotiation remains an important level, Belgian has a highly hierarchical collective negotiation structure and company collective negotiation is in some aspects overlooked with very few studies on this matter. Our study demonstrates the diversity of experiences across companies and underlies the necessity to investigate how different levels of negotiation affect workers. Second, few studies – and none for Belgium – have captured company collective negotiation in a quantitative way. Collecting data on social dialogue quality is complex and whilst this study uses an indicator of perceived change in social dialogue quality, we also demonstrate a relationship between the instruments implemented to protect workers within companies (e.g. risk assessment, role of the health and safety adviser) and social dialogue quality that shows that social dialogue is not a floating concept but an indicator of company practices. For instance, whilst the risk assessment is one of the preconditions for effective workers’ representation and consultation on health and safety (Walters & Nichols, 2006), the reverse was also true during the pandemic: when union health and safety representatives are present in the company, the risk assessment seems to be conducted more often (Cai et al., 2022). Further studies could investigate the causal nature of the relationship between the instruments implemented to protect the workforce and the quality of social negotiation. Finally, this study shows the empirical benefits of considering collective negotiation as a major cofounder in occupational health. Whilst the effects of work and employment characteristics are well documented in social epidemiology, the democratic nature of the workplace appears to be a very often forgotten level of public health. The COVID-19 pandemic might have exacerbated the importance of collective negotiation in protecting workers physical but also mental health and, whilst we are leaning towards a post-pandemic world, it could be hoped that these aspects will be taken into consideration in a more systematic way in further research.

## Supporting information

Supplementary file S2. Non-weigthed models

Supplementary file S1. Descriptive Statistics

## Data Availability

The dataset cannot be openly shared due to the collection of sensitive individual data and sample size. Specific data access on a restricted number of items can be formally requested to the authors.

## Glossary

95%CI: 95 per cent confidence interval
CPPT: Comité pour la Prévention et la Protection au Travail
CPPW: Comité voor Preventie en Bescherming op het Werk
EU: European Union
ILO: International Labour Organization
OR: Odds Ratio
OSH: Occupational Safety and Health
RR: Relative Risk

## Supplementary files

Supplementary file S1. Descriptive statistics

Supplementary file S2. Non-weighted models

## Footnotes

1 Companies belonging to critical sectors and essential services have been listed by the federal government on several occasions and initially in the annex to the Ministerial Order of 23 March 2020 on emergency measures to limit the spread of COVID-19.

2 The notions of home working and telework are used interchangaebly in this study even though they have different legal implications (International Labour Organization, 2020).

