## Supplementary file S2. Non-weigthed models for "Social dialogue quality and workers’ health as perceived by Belgian trade union representatives during the COVID-19 pandemic"

**Supplementary file S2. Descriptive statistics**

|  |  | Not weighted |  | Weighted |
| --- | --- | --- | --- | --- |
| Variables | | N=469. |  | N=469 |
| Pandemic physical health | | | | |
|  | 0 (good or average) | 366 (78.0%) |  | 370 (78.9%) |
|  | 1 (poor) | 103 (22.0%) |  | 99 (21.1%) |
| Pandemic mental health | | | | |
|  | 0 (good or average) | 263 (56.1%) |  | 268 (57.2%) |
|  | 1 (poor) | 206 (43.9%) |  | 201 (42.8%) |
| Quality of social dialogue since virus outbreak | | | | |
|  | Same | 257 (54.8%) |  | 249 (53.1%) |
|  | Better | 33 (7.0%) |  | 36 (7.8%) |
|  | Do not know | 38 (8.1%) |  | 42 (9.0%) |
|  | Worse | 141 (30.1%) |  | 141 (30.1%) |
| Sex composition | | | | |
|  | Mainly men | 94 (20.0%) |  | 91 (19.3%) |
|  | Half women | 255 (54.4%) |  | 258 (55.1%) |
|  | Mainly women | 117 (24.9%) |  | 118 (25.1%) |
| Manual or intellectual work | | | | |
|  | Both | 199 (42.4%) |  | 194 (41.5%) |
|  | Intellectual | 205 (43.7%) |  | 206 (44.0%) |
|  | Manual | 65 (13.9%) |  | 68 (14.6%) |
| Average age of the workforce | | | | |
|  | <40 | 102 (21.7%) |  | 98 (21.0%) |
|  | >40 | 158 (33.7%) |  | 162 (34.5%) |
|  | 40 | 209 (44.6%) |  | 209 (44.5%) |
| Company size | | | | |
|  | <50 | 40 (8.5%) |  | 46 (9.8%) |
|  | >1200 | 106 (22.6%) |  | 110 (23.4%) |
|  | 100-299 | 100 (21.3%) |  | 93 (19.9%) |
|  | 300-599 | 84 (17.9%) |  | 78 (16.7%) |
|  | 50-99 | 70 (14.9%) |  | 72 (15.4%) |
|  | 600-1200 | 69 (14.7%) |  | 70 (14.8%) |
| Langage of the questionnaire | | | | |
|  | French | 277 (59.1%) |  | 264 (56.3%) |
|  | Dutch | 192 (40.9%) |  | 205 (43.7%) |
| Pre-pandemic physical health | | | | |
|  | 0 (good or average) | 452 (96.4%) |  | 453 (96.6%) |
|  | 1 (poor) | 17 (3.6%) |  | 16 (3.4%) |
| Pre-pandemic mental health | | | | |
|  | 0 (good or average) | 426 (90.8%) |  | 428 (91.2%) |
|  | 1 (poor) | 43 (9.2%) |  | 41 (8.8%) |
| New collective agreement made during the pandemic | | | | |
|  | Yes | 132 (28.1%) |  | 123 (26.2%) |
|  | Do not know | 84 (17.9%) |  | 86 (18.3%) |
|  | No | 253 (53.9%) |  | 260 (55.5%) |
| Perceived role of the workplace physician | | | | |
|  | Average | 121 (25.8%) |  | 116 (24.6%) |
|  | Do not know | 52 (11.1%) |  | 57 (12.1%) |
|  | Good | 166 (35.4%) |  | 165 (35.2%) |
|  | Poor | 130 (27.7%) |  | 131 (28.0%) |
| Risk evaluation made during the pandemic | | | | |
|  | Yes | 178 (38.0%) |  | 176 (37.6%) |
|  | Do not know | 121 (25.8%) |  | 125 (26.8%) |
|  | No | 170 (36.2%) |  | 167 (35.6%) |
| Percentage working from home during the first lockdown (18 March to 3 May 2020) | | | | |
|  | 0 to 25 | 183 (39.0%) |  | 182 (38.9%) |
|  | 50 | 51 (10.9%) |  | 49 (10.5%) |
|  | 75 to 100 | 166 (35.4%) |  | 167 (35.7%) |
|  | Do not know | 69 (14.7%) |  | 70 (14.9%) |
| Percentage working from home after the first lockdown | | | | |
|  | 0 to 25 | 182 (38.8%) |  | 185 (39.5%) |
|  | 50 | 62 (13.2%) |  | 61 (13.0%) |
|  | 75 to 100 | 150 (32.0%) |  | 147 (31.3%) |
|  | Do not know | 75 (16.0%) |  | 76 (16.2%) |
| Social plan implemented during the pandemic | | | | |
|  | No | 417 (88.9%) |  | 421 (89.9%) |
|  | Do not know | 27 (5.8%) |  | 27 (5.7%) |
|  | Yes | 25 (5.3%) |  | 21 (4.5%) |
| Furlough during the first lockdown (18 March to 3 May 2020) | | | | |
|  | No | 229 (48.8%) |  | 240 (51.2%) |
|  | Do not know | 15 (3.2%) |  | 18 (3.7%) |
|  | Yes | 225 (48.0%) |  | 211 (45.0%) |
| Furlough after the first lockdown | | | | |
|  | No | 261 (55.7%) |  | 272 (58.1%) |
|  | Do not know | 27 (5.8%) |  | 30 (6.5%) |
|  | Yes | 181 (38.6%) |  | 166 (35.4%) |
| Work in a key sector | | | | |
|  | No | 149 (31.8%) |  | 145 (31.0%) |
|  | Do not know | 17 (3.6%) |  | 19 (4.1%) |
|  | Yes | 303 (64.6%) |  | 304 (64.9%) |
