## Supplementary file S1. Descriptive Statistics for "Social dialogue quality and workers’ health as perceived by Belgian trade union representatives during the COVID-19 pandemic"

**Supplementary file S3. Non-weighted models**

**Mental health (non-weighted)**

|  | No adjustment | | |  | Company adjustment | | |  | Pre-pandemic health adjustment | | |  | Full adjustment | | |
| --- | --- | --- | --- | --- | --- | --- | --- | --- | --- | --- | --- | --- | --- | --- | --- |
|  | RR | CI- | CI+ |  | RR | CI- | CI+ |  | RR | CI- | CI+ |  | RR | CI- | CI+ |
| (Intercept) | 0.38 | 0.33 | 0.45 |  | 0.35 | 0.21 | 0.61 |  | 0.33 | 0.19 | 0.58 |  | 0.28 | 0.15 | 0.53 |
| Dialogue: better | 1.19 | 0.8 | 1.79 |  | 1.25 | 0.85 | 1.85 |  | 1.26 | 0.85 | 1.86 |  | 1.2 | 0.8 | 1.79 |
| Dialogue: do not know | 1.1 | 0.74 | 1.65 |  | 1.2 | 0.79 | 1.82 |  | 1.27 | 0.84 | 1.91 |  | 1.47 | 0.97 | 2.22 |
| Dialogue: worse | 1.43 | 1.15 | 1.78 |  | 1.46 | 1.17 | 1.81 |  | 1.45 | 1.17 | 1.79 |  | 1.22 | 0.98 | 1.51 |
| Sex: Half women |  |  |  |  | 0.93 | 0.7 | 1.24 |  | 0.95 | 0.72 | 1.26 |  | 1.01 | 0.76 | 1.35 |
| Sex: Mainly women |  |  |  |  | 0.98 | 0.7 | 1.36 |  | 0.96 | 0.7 | 1.32 |  | 0.99 | 0.72 | 1.37 |
| Sex: do not know |  |  |  |  | 1.61 | 0.8 | 3.25 |  | 1.78 | 0.85 | 3.73 |  | 2.04 | 0.86 | 4.86 |
| age: >40 |  |  |  |  | 1.09 | 0.82 | 1.45 |  | 1.15 | 0.87 | 1.53 |  | 1.13 | 0.85 | 1.51 |
| age: 40 |  |  |  |  | 1.14 | 0.86 | 1.5 |  | 1.17 | 0.89 | 1.52 |  | 1.13 | 0.86 | 1.48 |
| size: >1200 |  |  |  |  | 1.46 | 0.96 | 2.2 |  | 1.3 | 0.87 | 1.96 |  | 1.46 | 0.96 | 2.22 |
| size: 100-299 |  |  |  |  | 1.12 | 0.72 | 1.75 |  | 1.01 | 0.66 | 1.55 |  | 1.05 | 0.68 | 1.62 |
| size: 300-599 |  |  |  |  | 1.15 | 0.73 | 1.8 |  | 1.03 | 0.67 | 1.61 |  | 1 | 0.65 | 1.55 |
| size: 50-99 |  |  |  |  | 1.23 | 0.78 | 1.94 |  | 1.12 | 0.72 | 1.74 |  | 1 | 0.65 | 1.53 |
| size: 600-1200 |  |  |  |  | 0.97 | 0.59 | 1.59 |  | 0.91 | 0.56 | 1.48 |  | 0.96 | 0.6 | 1.52 |
| Type of work: Intellectual |  |  |  |  | 0.77 | 0.61 | 0.97 |  | 0.78 | 0.62 | 0.98 |  | 0.81 | 0.61 | 1.08 |
| Type of work: Manual |  |  |  |  | 0.77 | 0.56 | 1.08 |  | 0.74 | 0.54 | 1.02 |  | 0.66 | 0.48 | 0.92 |
| Langage: NL |  |  |  |  | 1 | 0.8 | 1.25 |  | 1.05 | 0.84 | 1.31 |  | 1.07 | 0.85 | 1.34 |
| CP: 200-299 |  |  |  |  | 0.86 | 0.61 | 1.22 |  | 0.86 | 0.62 | 1.2 |  | 0.85 | 0.61 | 1.17 |
| CP: 300-399 |  |  |  |  | 1.17 | 0.84 | 1.63 |  | 1.11 | 0.8 | 1.53 |  | 0.94 | 0.68 | 1.3 |
| CP: None |  |  |  |  | 0.64 | 0.29 | 1.43 |  | 0.59 | 0.28 | 1.23 |  | 0.61 | 0.32 | 1.16 |
| CP: Public sector |  |  |  |  | 0.77 | 0.48 | 1.26 |  | 0.79 | 0.5 | 1.23 |  | 0.71 | 0.45 | 1.13 |
| Pre-pandemic mental health |  |  |  |  |  |  |  |  | 2.33 | 1.97 | 2.76 |  | 2.38 | 1.96 | 2.88 |
| New agreement: Do not know |  |  |  |  |  |  |  |  |  |  |  |  | 1.08 | 0.79 | 1.47 |
| New agreement: No |  |  |  |  |  |  |  |  |  |  |  |  | 0.99 | 0.78 | 1.26 |
| Role of physician: Do not know |  |  |  |  |  |  |  |  |  |  |  |  | 0.91 | 0.55 | 1.5 |
| Role of physician: Good |  |  |  |  |  |  |  |  |  |  |  |  | 0.99 | 0.73 | 1.34 |
| Role of physician: Poor |  |  |  |  |  |  |  |  |  |  |  |  | 1.61 | 1.23 | 2.09 |
| Risk evaluation: Do not know |  |  |  |  |  |  |  |  |  |  |  |  | 0.72 | 0.54 | 0.96 |
| Risk evaluation: No |  |  |  |  |  |  |  |  |  |  |  |  | 1.17 | 0.93 | 1.48 |
| Home working (t1): 50% |  |  |  |  |  |  |  |  |  |  |  |  | 0.93 | 0.59 | 1.45 |
| Home working (t1): 75 to 100% |  |  |  |  |  |  |  |  |  |  |  |  | 1.03 | 0.68 | 1.54 |
| Home working (t1): Do not know |  |  |  |  |  |  |  |  |  |  |  |  | 1.01 | 0.71 | 1.43 |
| Home working (t2): 50% |  |  |  |  |  |  |  |  |  |  |  |  | 1.19 | 0.77 | 1.82 |
| Home working (t2): 75 to 100% |  |  |  |  |  |  |  |  |  |  |  |  | 0.95 | 0.63 | 1.41 |
| Home working (t2): Do not know |  |  |  |  |  |  |  |  |  |  |  |  | 0.94 | 0.65 | 1.36 |
| Redundancy plan: Do not know |  |  |  |  |  |  |  |  |  |  |  |  | 0.83 | 0.48 | 1.42 |
| Redundancy plan: Yes |  |  |  |  |  |  |  |  |  |  |  |  | 1.36 | 0.96 | 1.93 |
| Furlough (t2): Do not know |  |  |  |  |  |  |  |  |  |  |  |  | 1.15 | 0.67 | 1.96 |
| Furlough (t2): Yes |  |  |  |  |  |  |  |  |  |  |  |  | 1.12 | 0.8 | 1.57 |
| Furlough (t1): Do not know |  |  |  |  |  |  |  |  |  |  |  |  | 0.26 | 0.09 | 0.76 |
| Furlough (t1): Yes |  |  |  |  |  |  |  |  |  |  |  |  | 0.88 | 0.63 | 1.24 |
| key worker: Do not know |  |  |  |  |  |  |  |  |  |  |  |  | 1.07 | 0.64 | 1.81 |
| key worker: Yes |  |  |  |  |  |  |  |  |  |  |  |  | 1.18 | 0.93 | 1.5 |

**Physical health (non-weighted)**

|  | No adjustment | | |  | Company adjustment | | |  | Pre-pandemic health adjustment | | |  | Full adjustment | | |
| --- | --- | --- | --- | --- | --- | --- | --- | --- | --- | --- | --- | --- | --- | --- | --- |
|  | RR | CI- | CI+ |  | RR | CI- | CI+ |  | RR | CI- | CI+ |  | RR | CI- | CI+ |
| (Intercept) | 0.19 | 0.15 | 0.25 |  | 0.18 | 0.07 | 0.48 |  | 0.17 | 0.06 | 0.43 |  | 0.2 | 0.07 | 0.58 |
| Dialogue: better | 1.09 | 0.54 | 2.2 |  | 1.06 | 0.58 | 1.92 |  | 0.75 | 0.4 | 1.41 |  | 0.57 | 0.27 | 1.21 |
| Dialogue: do not know | 1.08 | 0.56 | 2.1 |  | 1.18 | 0.59 | 2.35 |  | 1.23 | 0.64 | 2.37 |  | 1.26 | 0.62 | 2.56 |
| Dialogue: worse | 1.39 | 0.96 | 2 |  | 1.45 | 1.01 | 2.09 |  | 1.47 | 1.03 | 2.09 |  | 1.27 | 0.87 | 1.87 |
| Sex: Half women |  |  |  |  | 0.94 | 0.56 | 1.56 |  | 0.98 | 0.6 | 1.6 |  | 1.05 | 0.61 | 1.79 |
| Sex: Mainly women |  |  |  |  | 1.64 | 0.94 | 2.86 |  | 1.81 | 1.05 | 3.12 |  | 1.86 | 1.03 | 3.37 |
| Sex: do not know |  |  |  |  | 2.09 | 0.46 | 9.43 |  | 2.32 | 0.49 | 11 |  | 3.32 | 0.67 | 16.38 |
| age: >40 |  |  |  |  | 0.87 | 0.54 | 1.4 |  | 0.94 | 0.6 | 1.47 |  | 0.96 | 0.61 | 1.51 |
| age: 40 |  |  |  |  | 1.06 | 0.69 | 1.65 |  | 1.06 | 0.71 | 1.6 |  | 1.01 | 0.66 | 1.55 |
| size: >1200 |  |  |  |  | 0.92 | 0.5 | 1.7 |  | 0.95 | 0.53 | 1.71 |  | 0.76 | 0.39 | 1.46 |
| size: 100-299 |  |  |  |  | 0.86 | 0.44 | 1.68 |  | 0.91 | 0.48 | 1.73 |  | 0.85 | 0.44 | 1.62 |
| size: 300-599 |  |  |  |  | 1.07 | 0.57 | 2 |  | 1.05 | 0.58 | 1.93 |  | 0.87 | 0.46 | 1.66 |
| size: 50-99 |  |  |  |  | 0.9 | 0.45 | 1.82 |  | 0.91 | 0.47 | 1.76 |  | 0.74 | 0.37 | 1.48 |
| size: 600-1200 |  |  |  |  | 0.49 | 0.21 | 1.11 |  | 0.55 | 0.25 | 1.24 |  | 0.47 | 0.21 | 1.04 |
| Type of work: Intellectual |  |  |  |  | 0.71 | 0.47 | 1.07 |  | 0.73 | 0.49 | 1.09 |  | 1.02 | 0.6 | 1.75 |
| Type of work: Manual |  |  |  |  | 0.7 | 0.47 | 1.06 |  | 0.91 | 0.56 | 1.5 |  | 0.94 | 0.54 | 1.63 |
| Langage: NL |  |  |  |  | 1.02 | 0.6 | 1.73 |  | 0.73 | 0.49 | 1.09 |  | 0.7 | 0.46 | 1.05 |
| CP: 200-299 |  |  |  |  | 1.31 | 0.69 | 2.51 |  | 1.2 | 0.65 | 2.22 |  | 1.2 | 0.63 | 2.29 |
| CP: 300-399 |  |  |  |  | 1.58 | 0.85 | 2.94 |  | 1.29 | 0.72 | 2.33 |  | 1.07 | 0.55 | 2.08 |
| CP: None |  |  |  |  | 0.88 | 0.21 | 3.68 |  | 0.86 | 0.21 | 3.43 |  | 0.88 | 0.2 | 3.88 |
| CP: Public sector |  |  |  |  | 1.25 | 0.56 | 2.77 |  | 1.11 | 0.54 | 2.25 |  | 1.29 | 0.64 | 2.57 |
| Pre-pandemic mental health |  |  |  |  |  |  |  |  | 4.04 | 2.89 | 5.65 |  | 4.16 | 2.71 | 6.4 |
| New agreement: Do not know |  |  |  |  |  |  |  |  |  |  |  |  | 1.08 | 0.64 | 1.8 |
| New agreement: No |  |  |  |  |  |  |  |  |  |  |  |  | 0.69 | 0.46 | 1.03 |
| Role of physician: Do not know |  |  |  |  |  |  |  |  |  |  |  |  | 0.64 | 0.3 | 1.37 |
| Role of physician: Good |  |  |  |  |  |  |  |  |  |  |  |  | 0.82 | 0.48 | 1.38 |
| Role of physician: Poor |  |  |  |  |  |  |  |  |  |  |  |  | 1.29 | 0.82 | 2.03 |
| Risk evaluation: Do not know |  |  |  |  |  |  |  |  |  |  |  |  | 0.62 | 0.38 | 1.01 |
| Risk evaluation: No |  |  |  |  |  |  |  |  |  |  |  |  | 1.04 | 0.69 | 1.58 |
| Home working (t1): 50% |  |  |  |  |  |  |  |  |  |  |  |  | 0.91 | 0.42 | 2 |
| Home working (t1): 75 to 100% |  |  |  |  |  |  |  |  |  |  |  |  | 0.68 | 0.36 | 1.27 |
| Home working (t1): Do not know |  |  |  |  |  |  |  |  |  |  |  |  | 0.97 | 0.51 | 1.85 |
| Home working (t2): 50% |  |  |  |  |  |  |  |  |  |  |  |  | 0.98 | 0.49 | 1.95 |
| Home working (t2): 75 to 100% |  |  |  |  |  |  |  |  |  |  |  |  | 1.09 | 0.59 | 2.02 |
| Home working (t2): Do not know |  |  |  |  |  |  |  |  |  |  |  |  | 1.04 | 0.54 | 2 |
| Redundancy plan: Do not know |  |  |  |  |  |  |  |  |  |  |  |  | 1.09 | 0.48 | 2.49 |
| Redundancy plan: Yes |  |  |  |  |  |  |  |  |  |  |  |  | 1.02 | 0.53 | 2 |
| Furlough (t2): Do not know |  |  |  |  |  |  |  |  |  |  |  |  | 1.22 | 0.46 | 3.24 |
| Furlough (t2): Yes |  |  |  |  |  |  |  |  |  |  |  |  | 0.93 | 0.53 | 1.64 |
| Furlough (t1): Do not know |  |  |  |  |  |  |  |  |  |  |  |  | 0.36 | 0.07 | 1.88 |
| Furlough (t1): Yes |  |  |  |  |  |  |  |  |  |  |  |  | 1.33 | 0.76 | 2.35 |
| key worker: Do not know |  |  |  |  |  |  |  |  |  |  |  |  | 1.26 | 0.47 | 3.35 |
| key worker: Yes |  |  |  |  |  |  |  |  |  |  |  |  | 1.34 | 0.87 | 2.06 |
